# Establishing Cutoffs for Streptococcal Antibody Titers in Uganda: Implications for Rheumatic Fever Diagnosis

**DOI:** 10.1101/19010447

**Authors:** Emmy Okello, Meghna Murali, Joselyn Rwebembera, Jenifer Atala, Nada Harik, Gloria Kaudha, Samalie Kitooleko, Chris T Longenecker, Emma Ndagire, Isaac Otim Omara, Linda Mary Oyella, Tom Parks, Jafes Pulle, Craig Sable, Rachel Sarnacki, Elizabeth Stein, Meghan Zimmerman, Nicholas de Klerk, Jonathan Carapetis, Andrea Beaton

## Abstract

**Background:** Central to rheumatic fever (RF) diagnosis is evidence of streptococcal exposure, specifically antistreptolysin O (ASO) and antideoxyribonuclease B (ADB) antibodies. It is unknown if these antibody titers should be adjusted to the background exposure rates of GAS or if published standards should be used. Here, we establish the normal values of ASO and ADB in Uganda and examine RF case detection using published vs. population-specific thresholds.

**Methods:** Participants (age 0-50 years) were recruited. ASO was measured in-country by nephelometric technique. ADB samples were sent to Australia (PathWest) for ADB determination by enzyme inhibition assay, andthe 80% upper limit values by age were established. The published standard values for ASO (200IU/ml) and ADB (375IU/ml) were compared to the Ugandan 80% upper limit of normal values (ULN) for RF case detection in children 5-15 years.

**Findings:** Of the 428 participants, 16 were excluded from analysis (9 sore throat, 1 skin sores, 5 fever, 4 echocardiograms showing occult RHD), and 183 of the remaining were children 5-15 years. The median ASO titer in this age group was 220 IU/ml, with the 80th percentile value of 389 IU/ml. The median ADB titer in this age group was 375 IU/ml, with the 80th percentile value of 568 IU/ml. Application of new Ugandan cutoffs to 528 children enrolled in our prospective RF study, reduced the number of definite RF cases to 120/528 (22·7%), as compared to 173/528 (32·8%) using published normal values.

**Interpretation:** The 80th percentile ULN for ASO and ADB are higher in Uganda than in other countries. Applying these higher values to RF diagnosis in Uganda results in higher diagnostic specificity, but some unknown loss in sensitivity. Implications of over-diagnosis and missed cases will be explored through a longitudinal follow-up study of children in the RF research program.

**Funding:** This work was supported by American Heart Association Grant #17SFRN33670607 / Andrea Beaton / 2017 and DELTAS Africa Initiative.

**Research in context:** *Evidence before this study:* We searched PubMed for data on normal values of streptococcal antibody titers within diverse populations between database inception and January 1, 2019, using the search terms (rheumatic fever) OR (streptococcal antibodies). Nine studies were identified, but only one was from sub-Saharan Africa (2018, Ethiopia) and it was limited by vague exclusion criteria and lack of data on anti-DNase B. Given the high burden of rheumatic heart disease in sub-Saharan Africa, further data is needed to determine normal streptococcal antibody titers in this population and to assess the clinical impact of different cutoffs for RF diagnosis.

*Added value of this study:* Our study utilized a rigorous approach to exclude patients with history of recent possible streptococcal exposure including skin and throat infection and employed echocardiography to exclude patients with pre-existing rheumatic heart disease. Additionally, this study was conducted in parallel to a larger epidemiological cohort study of rheumatic fever in Uganda, allowing us, for the first time, to prospectively determine how utilization of different streptococcal antibody titer cutoffs affect diagnosis of rheumatic fever.

*Implications of all the available evidence:* Rheumatic fever remains a challenging diagnosis based on a clinical decision rule with imperfect sensitivity and specificity. Improved understanding of streptococcal antibody titers in rheumatic heart disease endemic populations may improve diagnostic performance. Our study also points to the need for development of a rheumatic fever diagnostic test, in order to provide a more definitive assessment of risk.

## Background

Improving the diagnosis of rheumatic fever (RF) in low-resource settings is imperative. Undiagnosed RF results in a missed opportunity to offer prophylaxis against recurrent group A streptococcal (Strep A) infections, the driver of RF and later rheumatic heart disease (RHD).^1^ The diagnosis of RF typically requires evidence of preceding Strep A infection.^2^ This evidence can take the form of a positive rapid antigen test or culture, but more commonly requires serum streptococcal antibody testing to detect evidence of Strep A infection prior to RF development.^2^ The two most common commercially available streptococcal antibody assays are anti-streptolysin O (ASO) titer and anti-DNase B (ADB), but a variety of interpretations exist regarding what constitutes elevated titers.^3^

The 2015 Jones criteria, the most widely applied criteria for RF diagnosis, recommend assessing acute and convalescent ASO and ADB antibody titers, with a two-fold increase over 2-4 weeks,^4^ as the most robust evidence of recent Strep A infection. However, in low resource settings, requiring multiple tests can be financially prohibitive and risks patients being lost to follow-up as antibody assays, if available at all, are usually restricted to major urban centers. Therefore, comparing a single titer to the upper limit of normal (ULN) is accepted as sufficient evidence of recent streptococcal infection.^5^ The ULN for streptococcal serology is determined by the 80^th^ percentile of a normally distributed curve of healthy population titers.^6^

The 80^th^ percentile cutoffs for ASOT and ADB vary geographically, likely reflecting differences in background Strep A exposure and potentially genetic variability in immune response. While investigations of population-based normal values have been conducted in North America,^7^ Asia,^8-10^ the Middle East,^3,11^ and the Pacific,^12^ there is limited data for ULN values in Africa, and no data from East Africa. The sensitivity and specificity of RF diagnosis will logically be dependent on the accepted ULN for streptococcal antibody titers within a population, however the size of this effect has not been studied.

In the context of a larger prospective study to determine the incidence of RF, the current study determined the ULN for ASO and ADB for children and adults in Uganda, utilizing a parametric regression modeling technique, as previously described.^5^ We subsequently evaluated the number of children diagnosed with RF, as part of the aforementioned ongoing epidemiological study, using the newly derived Ugandan population ULN values (both as a standardized single value for children 5-14 and as age-specific values by year) compared to commonly used normal ULN values, and ULN values established through similar methods in Fiji.^5^

## Materials and Methods

### Setting

Uganda is a low-income country in East Africa home to approximately 44 million people and ranked 162 of 189 nations on the United Nations Development Program’s 2017 Human Development Index.^13^ Approximately 84% of the population lives in rural areas, and 48% of the population is less than 15 years of age. The national referral hospital, Mulago National Referral Hospital, is located in the capital city, Kampala and includes the Uganda Heart Institute, which provides tertiary cardiac services, including cardiac surgery and interventional cardiac catheterization, for the country. In Uganda, 16% of children are colonized with Strep A^14^ and Strep A pharyngitis is common with an incidence of 10·3 cases per 100 child weeks *(pilot data* American Heart Association funded RF epidemiological surveillance study). Approximately 2·5% of Ugandan school-going children show evidence of RHD by echocardiographic screening^14-16^. Clinical RHD is diagnosed late, with 85% of patients diagnosed only at the time of advanced cardiac disease and complications.^17^ The average age of death from RHD in Uganda is 29 years^18^.

### Determining ULN values for ASO and ADB

Recruitment for the ULN values study was achieved through advertisement at several primary schools in Kampala and by direct invitation of children (>1 year) and adults (≤ 50 years) accompanying siblings/parents to the Uganda Heart Institute. Recruitment targeted 400 participants: 100 participants in the two age strata known to have the highest background antistreptococcal antibody titers (5-10 years and 10-15 years) and 50 participants from all other age strata (1-4 years, 15-24 years, 25-34 years, 35-50 years). Participants were prospectively screened for current or recent Strep A infection. We excluded any participant who reported a history of RF, RHD, or invasive Strep A disease or history of sore throat or skin sores in the past 14 days. We also excluded participants who had a temperature of >38·0°C on the day of enrollment. All participants underwent focused echocardiography by a cardiologist, with review of the left-sided cardiac valves. Those showing evidence of RHD according to the 2012 WHF criteria^19^ were also excluded.

### Confirming Rheumatic Fever

In parallel to the ULN values study, participants aged 3-18 with signs/symptoms of possible RF were being recruited in Lira District, Northern Uganda as part of an American Heart Association funded RF epidemiological surveillance study. Inclusion criteria for the epidemiological surveillance study required children to have one of three presentations (1) fever and joint pains (2) suspicion of acute rheumatic carditis or (3) suspicion of Sydenham’s chorea. Children with a known alternate diagnosis responsible for the required presentation (e.g. sickle cell crisis) were excluded from the study. Enrolled children had full work-up for RF including clinical examination and history, rapid GAS testing, throat culture, ASO and ADB titers, limited echocardiography, ECG. Testing for common alternate diagnoses was also employed with rapid malaria testing, blood smear, influenza PCR, and auxiliary testing for clinical suspicion of other conditions (i.e. blood culture for suspected typhoid fever, or joint aspirate and culture for suspected septic arthritis). Diagnosis of RF was made by remote chart review and telemedicine review of ECG and echocardiogram according to the 2015 Jones Critiera^2^ employing four streptococcal titer cut offs: 80% ULN determined by this study (both age-stratified and as a single population value), commonly used standard reference values (ASO titer ≥ 200 IU/mL and ADB ≥ 310 IU/mL), and robust population data from Fiji (ASO ≥ 276 IU/mL and ADB ≥ 499 IU/mL).

### Sample collection, transport, and testing

Blood specimens were obtained by standard venipuncture and placed immediately in a cool box. Samples were transported to the laboratory on the day of collection. Upon arrival at the laboratory, the samples were centrifuged, and the serum was divided into two aliquots. One aliquot was used to determine the ASO titer at MBN Laboratories in Uganda. and the second aliquot was stored at −80°C. After all samples had been collected, the frozen aliquots were shipped on dry ice to Perth, Australia for ADB titer determination at PathWestLaboratories.

ASO titers were measured by nephelometric technique (Beckman Coulter, Fullerton, CA) as described previously^5^. ADB titers were measured by enzyme inhibition assay (bioMerieux, Marcy l’Etoile, France), as described previously^5^. Both methods provide an inexact figure for low titers (titers of <60 IU/ml for ASO and titers of <100 IU/ml for ADB); for these values, we estimated mid-titer values (a titer of 30 IU/ml for ASO and a titer of 50 IU/ml for ADB), as previously described^5^.

### Statistical Methods

#### Determining ULN values for ASO and ADB

We used an amended version of previously described techniques for constructing age-specific ASO and ADB titer reference ranges^5^. We followed the procedures described by Wright and Royston^20^ using the Stata routine *xriml*. The raw data for both ASO and ADB titers were transformed to normality and then fitted to regression curves using fractional polynomials to describe the non-linearity of the distributions with age. From the resulting fitted curves, we obtained median and 80% upper-limit-of-normal values for five age groups (1-4 y, 5-14 y, 15-24 y, 25-34 y, 35-50 y) and also by year of age for children aged 5 to 14 years. Data were analyzed using Stata 15.

#### Confirming Rheumatic Fever

The number and percentage of subjects with evidence of recent Streptococcal infection by rapid Strep A testing, throat culture, ASO titer, and ADB titer, and a diagnosis of RF were determined for each of our four streptococcal titer cut off groups. Fisher’s Exact Test was used to compare the proportion of children diagnosed with definite RF using Ugandan normal values by yearly age to Ugandan standardized single values, commonly used normal values, and population normal values from Fiji.

#### Ethical Approval

Institutional review board (IRB) approval was obtained from the Research and Ethics Committee at Makerere University School of Medicine, the Children’s National Hospital IRB, and the Uganda National Council for Science and Technology IRB. We approached all participants for their consent, and the study was explained in detail by a research nurse in the participants’ primary language. All participants provided written informed consent before information was collected and children were enrolled only if written consent was obtained from a parent or guardian and written assent by children aged 8 years or older.

## Results

### Determining ULN values for ASO and ADB

In total, 428 participants were recruited of which 250 (58%) were female. Study participants included 56 children 1-4 years of age, 90 children 5-9 years of age, 102 children 10-14 years of age, 70 young adults 15-24 years of age, 52 adults 25-34 years of age, and 52 adults 35-50 years of age, and 6 individuals with incomplete age data recorded. Of these 428 participants, 22 (5%) were excluded: six for incomplete age data, nine for sore throat in the last 14 days, one for skin sores in the last 14 days, two for fever on the day of enrollment, and four for echocardiographic evidence of RHD. Additionally, 16 ASO titer samples and 12 ADB titer samples were not of sufficient quantity for processing, leaving 390 samples for ASO analysis and 394 samples for ADB analysis.

When the data were log-transformed, there was one ADB value that was noted to be an extreme outlier (2640 IU/mL) and this value was removed. There were no extreme outliers for ASO and all values were retained for analysis.

### Normal ASO and ADB Population Titers

There was a peak in the mean titers of both ASO and ADB in the 5 to 14-year-old age group, with a gradual decrease occurring following this peak (Appendix 3 and 4). The estimated median and 80% upper-limit-of-normal values for five age groups are presented in Appendix 1. Appendix 2 presents these data in more detail for children aged 5 to 14 years of age. The estimated Ugandan pediatric population standardized 80% upper-limit-of-normal ASO and ADB titers were 389 IU/ml and 568 IU/ml respectively.

#### Confirming Rheumatic Fever

At the time of this analysis, there were 225 children enrolled in the RF epidemiological survey; 192 of whom had complete streptococcal antibody titer data available for review. The median age of participants was 10 years (IQR: 7), and 51·5% of participants were female. Utilizing population-based normal ASO and ADB values by yearly age for Uganda obtained from this study, 45 (23·4%) participants were diagnosed with RF, compared to 44 (22·9%, p=1·0) participants when a single standard value for Uganda was utilized, 64 (33·3%, p=0·04) participants when commonly used normal values were utilized, and 53 (27·6%, p=0·4) participants when population normal values for Fiji were utilized (Appendix 5).

## Discussion

We found the normal values for streptococcal serology in Uganda to be higher than those reported in many other countries (Appendix 6-7).

Strep A serological values, as seen in our study, consistently vary by age, with peak values in children 5-14 years old.^5,12,21,23^ Serological values have also been noted to vary by time of year in places with varied seasonal temperatures.^3,12^ Both of these factors reflect differences in streptococcal exposure, not inherent differences in immunological response between populations. Differences in serological values attributable to Strep A exposure is further confirmed by high ADB titers in Fiji reflecting high rates of Strep A skin infection (more likely to cause an ADB response) compared to Strep A sore throat, and possibly both the longer duration of ADB as compared to ASOT response and the potential for cumulative serological responses where multiple infections are likely in the same person over a short time.^5^

This study addresses the relevant and novel question of the impact of utilizing population-specific streptococcal titers, as compared to more general cutoffs, potentially generated in populations with lower background streptococcal exposure, to make the diagnosis of RF. Our data suggest that using population-specific titers developed by yearly age, as compared to a single elevated population-specific titer level for those 5-14 years of age, had little impact on the diagnosis of RF as only one case no longer met criteria for RF. However, utilizing population-based normal values from Fiji, where Strep A pharyngitis is less common, resulted in a non-significant but notable additional eight cases of RF (20% increase in diagnosed cases), and utilizing commonly used standard laboratory reference ranges resulted in a significant increase in the diagnosis of RF with 20 additional cases (45% increase in diagnosed cases). Further phenotypic studies are needed to evaluate if Ugandan children diagnosed with RF with lower ASO and ADB titers show evidence of carditis, the most important and lasting manifestation of RF, at diagnosis or during longitudinal follow-up, or if they have increased risk of recurrent RF. A study addressing these questions is currently ongoing in Uganda.

As ASO and ADB titers can remain elevated for many months after Strep A infection, despite efforts to exclude participants with a recent history of sore throat or impetigo, we are likely to have included some children whose titers were in the process of returning to baseline level after a Strep A infection. While the annual incidence of Strep A pharyngitis and impetigo remains to be definitively determined in Uganda, pilot data from Gulu (Northern Uganda) found 41% of children experienced a Strep A positive sore throat over four weeks of active surveillance, translating to 10·3 Strep A positive sore throats per 100 child-weeks (unpublished). With this number of children experiencing Strep A sore throat, population titers for unexposed children may be difficult to determine. Children receiving penicillin or cotrimoxazole prophylaxis, which would include young children with sickle cell disease, children with renal disease, those living with HIV, and those on secondary prophylaxis for RHD, could potentially be used as a population for Strep A unexposed studies. However, using these populations of children would introduce additional confounders.

Our study provides the ULN normal values for streptococcal serology for children and adults in Uganda and demonstrates that using a single population based cutoff, as compared to normal values by year of age, for normal vs. abnormal titers is appropriate for children aged 5-14 years of age. Perhaps more importantly, these data also highlight that currently available tests for serological confirmation of recent Strep A infection lack precision, in particular in populations with high background rates of Strep A infection. There is a need for better tests that truly reflect recent infection rather than those that have occurred in the past months or even years. In the meantime though we consider the appropriateness of these cutoffs as compared to international standardized cutoffs to be an open question, these ULN values provide some guidance to clinicians working in Uganda. Further longitudinal investigations will reveal if using population-specific cutoffs generated from Strep A endemic environments appropriately improves the specificity or inappropriately lowers the sensitivity of RF diagnosis.

### Data Sharing

Study data will not be publicly available. Any data requests should be made to the corresponding author.

## Acknowledgements

This work was supported by American Heart Association Grant #17SFRN33670607 / Andrea Beaton / 2017 and by DELTAS Africa Initiative. The investigators would like to thank General Electric Healthcare for providing echocardiography equipment and all the study participants and staff members who participated in and assisted with this study.

